# Ultra-High-Resolution CT Follow-Up in Patients with Imported Early-Stage Coronavirus Disease 2019 (COVID-19) Related Pneumonia

**DOI:** 10.1101/2020.03.31.20048256

**Authors:** Yu Lin, Shaomao Lv, Jinan Wang, Jianghe Kang, Youbin Zhang, Zhipeng Feng

## Abstract

**Background:** An ongoing outbreak of mystery pneumonia in Wuhan was caused by coronavirus disease 2019 (COVID-19). The infectious disease has spread globally and become a major threat to public health.

**Purpose:** We aim to investigate the ultra-high-resolution CT (UHR-CT) findings of imported COVID-19 related pneumonia from the initial diagnosis to early-phase follow-up.

**Methods:** This retrospective study included confirmed cases with early-stage COVID-19 related pneumonia imported from the epicenter. Initial and early-phase follow-up UHR-CT scans (within 5 days) were reviewed for characterizing the radiological findings. The normalized total volumes of ground-glass opacities (GGOs) and consolidations were calculated and compared during the radiological follow-up by artificial-intelligence-based methods.

**Results:** Eleven patients (3 males and 8 females, aged 32-74 years) with confirmed COVID-19 were evaluated. Subpleural GGOs with inter/intralobular septal thickening were typical imaging findings. Other diagnostic CT features included distinct margins (8/11, 73%), pleural retraction or thickening (7/11, 64%), intralesional vasodilatation (6/11, 55%). Normalized volumes of pulmonary GGOs (*p*=0.003) and consolidations (*p*=0.003) significantly increased during the CT follow-up.

**Conclusions:** The abnormalities of GGOs with peripleural distribution, consolidated areas, septal thickening, pleural involvement and intralesional vasodilatation on UHR-CT indicate the diagnosis of COVID-19. COVID-19 cases could manifest significantly progressed GGOs and consolidations with increased volume during the early-phase CT follow-up.

## 1. Introduction

An outbreak of mystery pneumonia suggesting inter-human transmission has been reported in Wuhan, China since December 2019 [1]. The novel infectious disease, currently known as coronavirus disease 2019 (COVID-19), has spread rapidly and become a global pandemic [1,2]. Severe acute respiratory syndrome (SARS) was the first emerging infectious disease of the new century caused by coronavirus with a tremendous threat and a high case-fatality rate of approximately 10% [3]. As another respiratory tract contagious disease caused by coronaviruses, COVID-19 may also lead to progressive respiratory failure and varied ground-glass opacity (GGO) pattern on chest CT as previously reported [3-7].

Positive CT findings could be found earlier than available results of real-time reverse-transcription-polymerase-chain-reaction (rRT-PCR) for patients with COVID-19 [8,9]. A considerable number of cases with early-stage COVID-19 demonstrated mild nonspecific symptoms (such as fever, fatigue and dry cough). Early-stage patients with mild symptoms may represent considerably infectivity and develop rapidly in a short period of time, which may lead to delay of medical treatment and expansion of epidemic situation [2,10]. Based on current clinical experience, the application of CT examination is helpful for preclinical screening, early diagnosis, timely isolation and initial follow-up for patients with high clinical suspicion of COVID-19 [9].

However, radiographic features of COVID-19 related pneumonia have been described in only few preliminary studies. Thus, we aim to determine the characteristics of imported early-stage COVID-19 on thin-section ultra-high-resolution CT (UHR-CT) images, and to evaluate the development of pulmonary lesions during a short-term CT follow-up.

## 2. Materials and Methods

### 2.1 Patients

This study was approved by our institutional review board; informed consent was waived for the retrospective nature. Records for patients with highly suspected COVID-19 from 20 January 2020 to 15 February 2020 in our hospital were reviewed. Our inclusion criteria were: (1) positive detection of COVID-19 by rRT-PCR; (2) necessary clinical and laboratory information; (3) available base-line UHR-CT scan; (4) initial UHR-CT scan with evidence of pneumonia (could be a base-line CT scan or a subsequent repeated CT scan); (5) another follow-up UHR-CT scan within 5 days. Patients with other causes of pneumonia (common bacterial and viral pathogens) were excluded

### 2.2 CT Imaging

UHR-CT scans were uniformly performed using a multi-detector spiral CT scanner (Ingenuity, Philips Medical Systems, the Netherlands) with a breath-hold after full inspiration. The CT protocols were as follows: tube voltage: 120 kV; automatic tube current; collimation: 64×0.625 mm; pitch: 1.2; matrix: 512×512 (for routine workup) or 1024×1024 (for further analysis); reconstruction technique: hybrid iterative algorithm (iDose^4^); thickness: 1 mm; increment: 1 mm. Multiplanar reconstruction, minimum intensity projection, and volume rendering were conducted on a professional workstation.

### 2.3 Data Processing

Two radiologists (with 20 and 6 years of experience of cardiothoracic radiology, respectively) reviewed all the UHR-CT images independently. The senior radiologist made the final decision when there was a discrepancy.

UHR-CT images of all cases were assessed for the following abnormalities (1) affected lobes; (2) distribution (peribronchovascular, peripleural and scattered/diffuse); (3) margin (sharp or indistinct); (4) specific signs (air bronchogram, intralobular interstitial thickening and/or interlobular septal thickening, intralesional vasodilatation, pleural retraction/thickening, and pleural effusion).

Quantitative analyses were automatically performed in all cases using an updated artificial intelligence-based image analysis system (Intelligent Evaluation System of Chest CT, Yitu Healthcare, China, https://www.yitutech.com/en). Pulmonary inflammatory lesions of the initial and followed-up UHR-CT images were intelligently recognized based on morphological features, and segmented based on the attenuation differences (areas of GGO and consolidation with a cut-off value of 250Hu, Figure 1).

**Fig 1.**
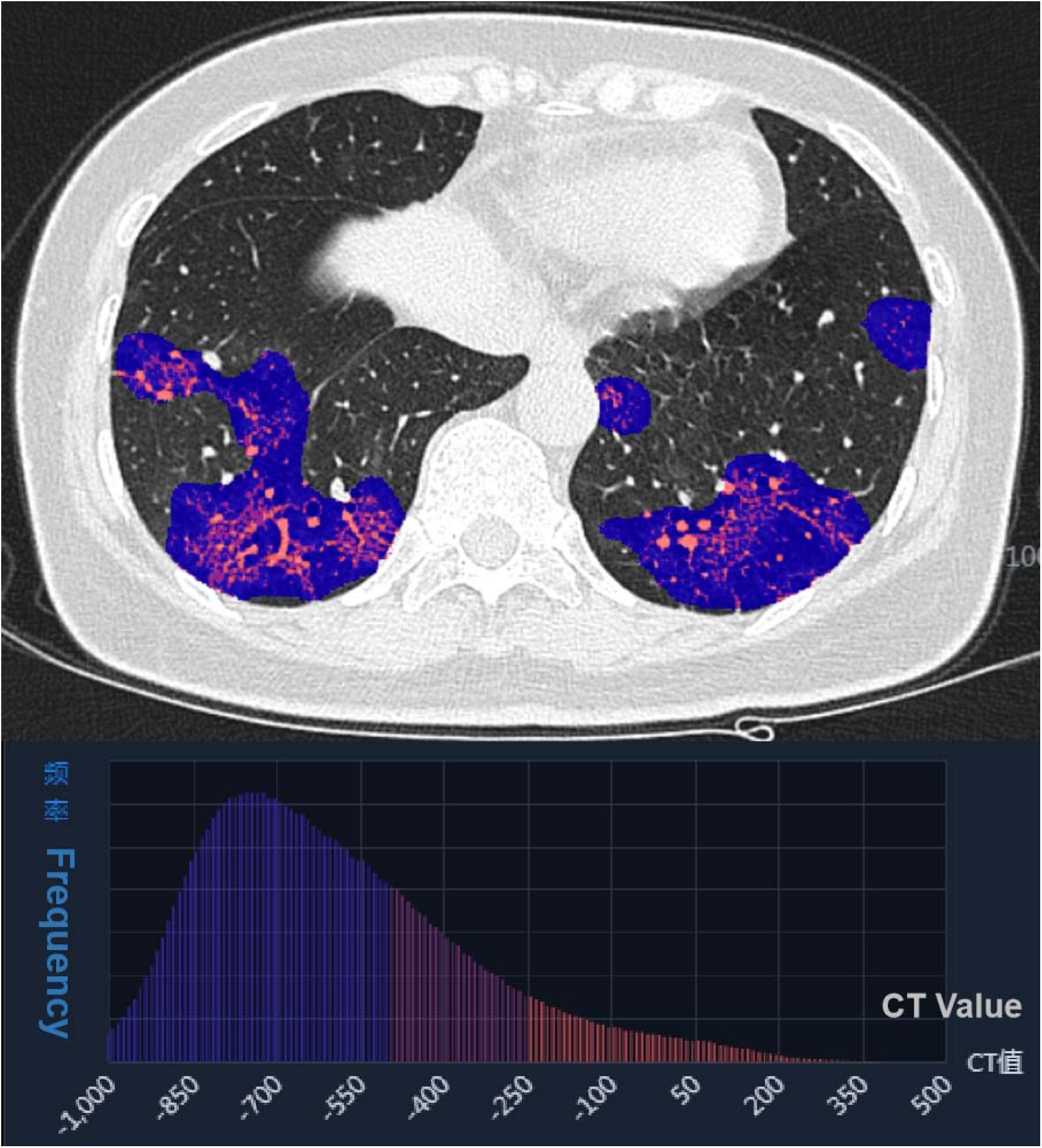
A heatmap of pulmonary inflammatory lesions on CT scan in the artificial-intelligence-based quantitative analyses. Red areas represent consolidation; blue and purple areas represent ground-glass opacity.

The estimated total volumes of different components of lesions (GGO and consolidation) were calculated automatically. Normalized volume (nV) was obtained by normalizing to the estimated total lung volume based on the following formula: nV= volume _GGO or consolidation_ / volume _lung_ × 100%.

### 2.4 Statistical Analysis

Statistical analyses were performed using Statistical Package for the Social Sciences (IBM Inc., USA). All quantitative data were presented as mean ± standard deviation or median (range). The comparisons of paired quantitative data during the CT follow-up were evaluated by the Wilcoxon test. A *p*-value of < 0.05 was defined as statistical significance.

## 3. Results

### 3.1 Clinical features

A total of 11 patients (3 males and 8 females, median age 53 years, range 32-74 years) with rRT-PCR confirmed COVID-19 related pneumonia were enrolled in the study (Table 1). All patients have a history of epidemiological exposure (8 cases had lived in or traveled to Wuhan, 3 cases had contact with confirmed cases from Wuhan).

**Table 1.** Demographic and clinical features of patients with COVID-19 (n=11)

| **Parameter** | **Number (%)** |
| --- | --- |
| **Gender (male/female)** | 3 (27%) / 8 (73%) |
| **Age (year old)** | 53 (32-74) |
| **Exposure history** |  |
| Recent Wuhan contact | 8 (73%) |
| Exposure to infected case | 3 (27%) |
| **Initial symptoms** |  |
| Fever | 9 (82%) |
| Cough | 6 (55%) |
| Fatigue | 4 (36%) |
| Stuffy and runny nose | 2 (18%) |
| Headache and dizziness | 1 (9%) |
| Muscle soreness | 1 (9%) |
| **Comorbidity** | 3 (27%) |
| **Laboratory findings** |  |
| Reduced leukocyte count | 3 (27%) |
| Reduced neutrophil count | 1 (9%) |
| Reduced lymphocyte count | 8 (73%) |
| Elevated C-reactive protein | 6 (55%) |

The most common symptoms were fever (9/11, 82%) in our case series. Mild to moderate symptoms of cough (6/11, 55%) and fatigue (4/11, 36%) were found in COVID-19 patients. Decreased lymphocyte count (8/11, 73%) and elevated C-reactive protein level (6/11, 55%) were major abnormal laboratory findings. Only three (27%) patients had comorbidities (diabetes, hyperuricemia, tuberculosis and cardiac disease). In patients with imported COVID-19 in our cohort, no severe cases (respiratory rate >30 breaths/min and/or requirement for mechanical ventilation) were found during the initial follow-up.

The base-line CT scan [obtained 3 (0.5-6) days after onset of clinical symptoms] showed positive results of pneumonia in 8 (73%) cases with COVID-19. In one case, the initial positive result of pneumonia was found in the third CT examination (6 days from symptoms onset) after hospitalization. Interestingly, all cases except one showed signs of pneumonia on CT images before achieving positive rRT-PCR results.

### 3.2 Initial CT findings

All of the eleven patients underwent at least two chest CT scans (median interval: 3 days, range: 2-5 days). Data from initial and follow-up chest CT imaging with positive findings (n=11) in patients with COVID-19 were presented in Tables 2 and 3, and Figure 2 and 3. There were 10 (91%) patients with peripleural distribution, 7 (64%) patients with peribronchovascular distribution, 9 (82%) patients with bilateral involvement, 9 (82%) patients with affected lower lobes, and 7 (64%) patients with multiple (≥3) involved lobes.

**Table 2.** Initial ultra-high-resolution CT findings of COVID-19 cases (n=11)

| **Findings** | **Patients** |
| --- | --- |
| **No. of Affected lobe(s)** |  |
| 1 | 2 (18%) |
| 2 | 2 (18%) |
| 3 - 5 | 7 (64%) |
| **Distribution** |  |
| Peripleural | 10 (91%) |
| Peribronchovascular | 7 (64%) |
| Scattered or diffuse | 3 (27%) |
| **Sharp margin** | 8 (73%) |
| **Radiologic signs** |  |
| Air bronchogram | 11 (100%) |
| Inter/intralobular<br>septal thickening | 11 (100%) |
| Pural retraction or thickening | 7 (64%) |
| Vasodilatation | 6 (55%) |
| Pleural effusion | 1 (9%) |

**Table 3.**
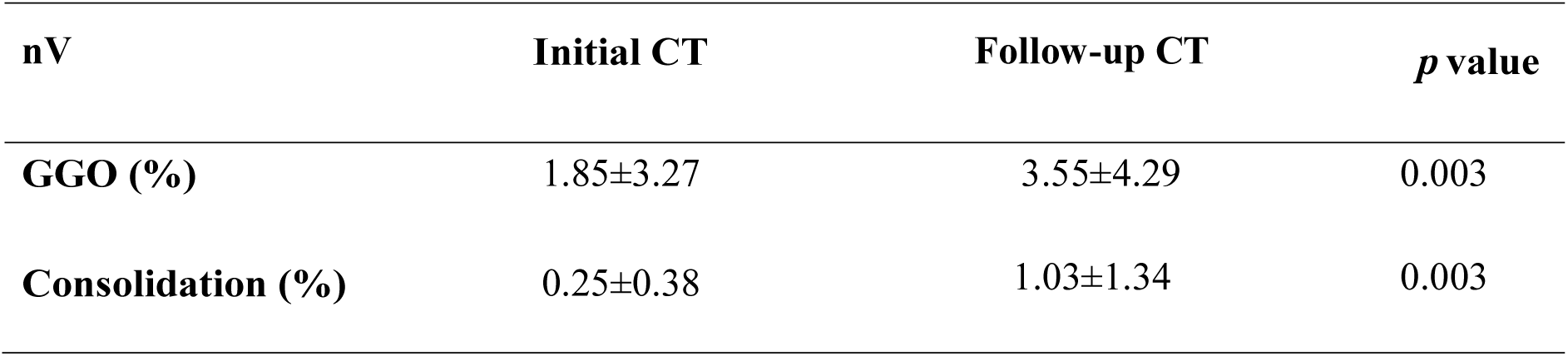
Comparison of normalized volume (nV) of ground-glass opacity (GGO) and consolidation during the CT follow-up (n=11)

**Fig 2.**
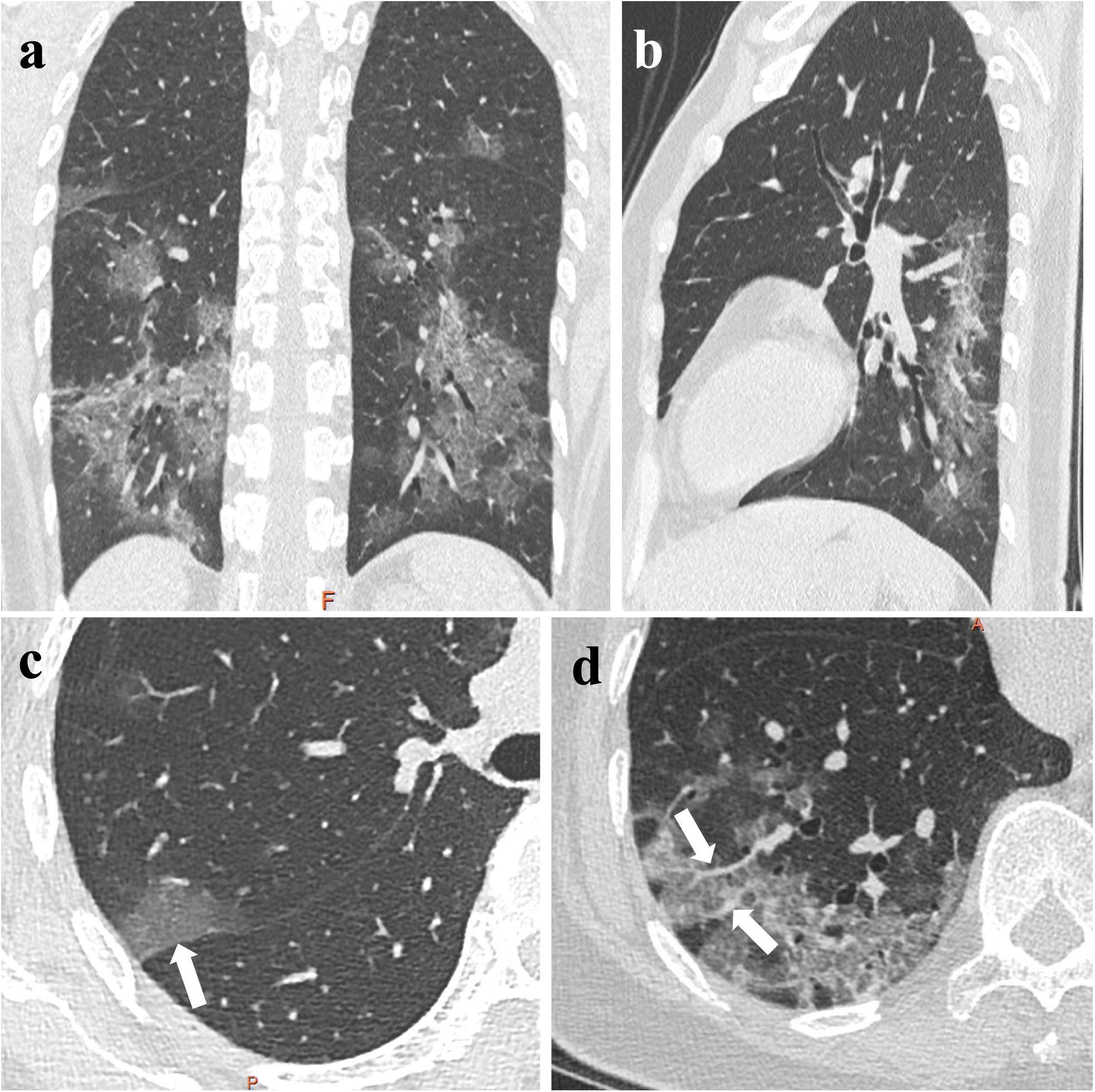
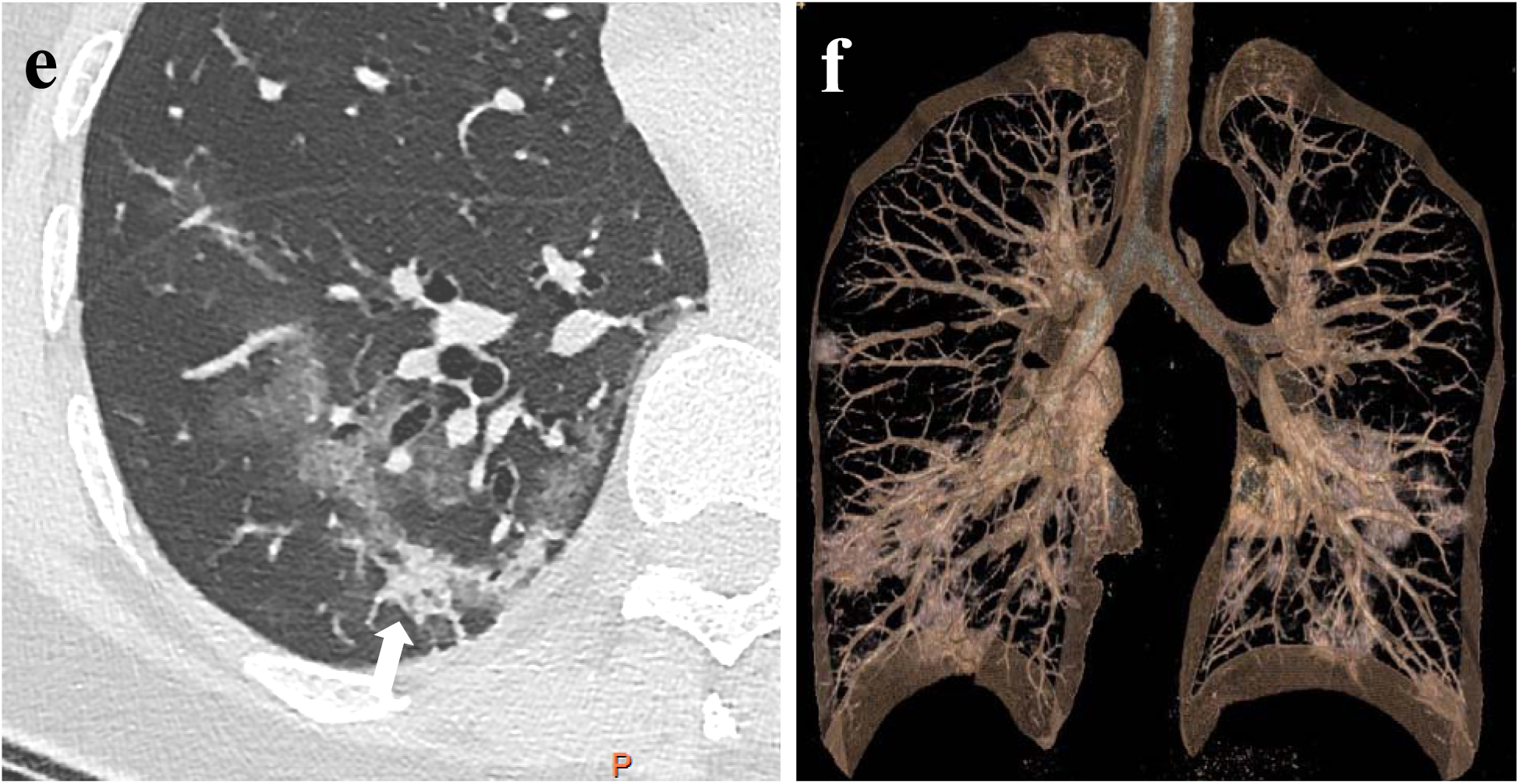
Initial coronal (a) and sagittal (b) CT images of a 53-year-old female with COVID-19 show multiple well-defined pulmonary lesions with subpleural and peribronchovascular distribution. The lesions show ground-glass opacity (GGO) with pleural retraction and thickening (arrow) (c), GGO with interlobular septal thickening and vasodilatation (arrow) (d), and GGO with patchy consolidation (arrow) and pleural retraction (e). Volume rendering image (f) vividly illustrates the lesion characteristics.

**Fig 3.**
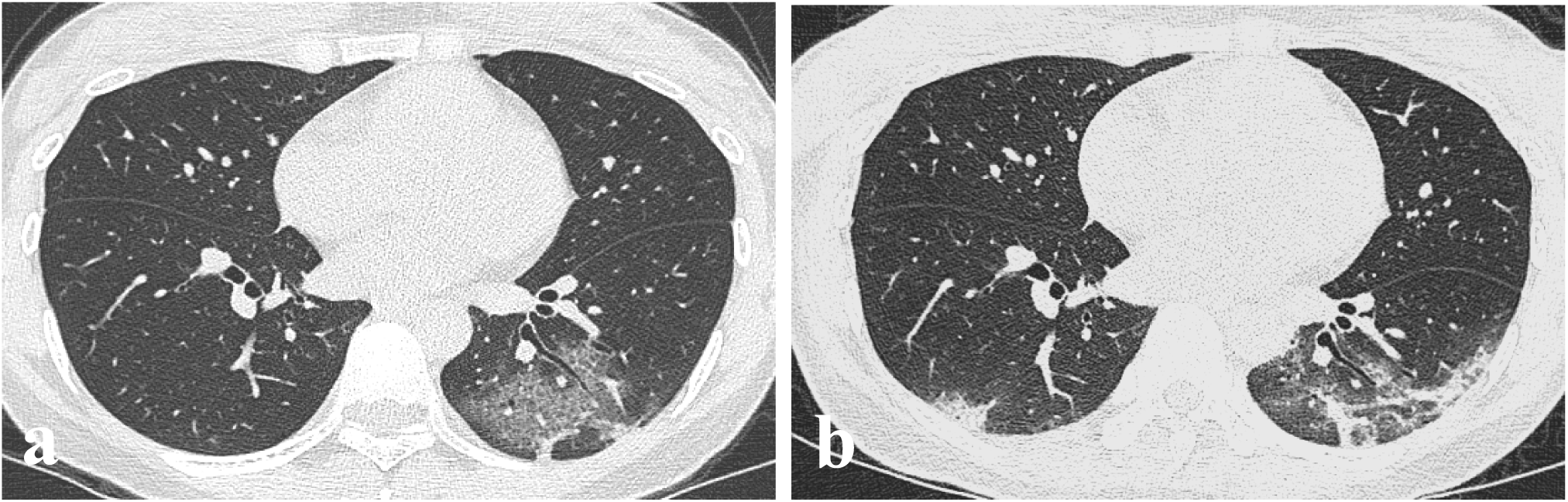
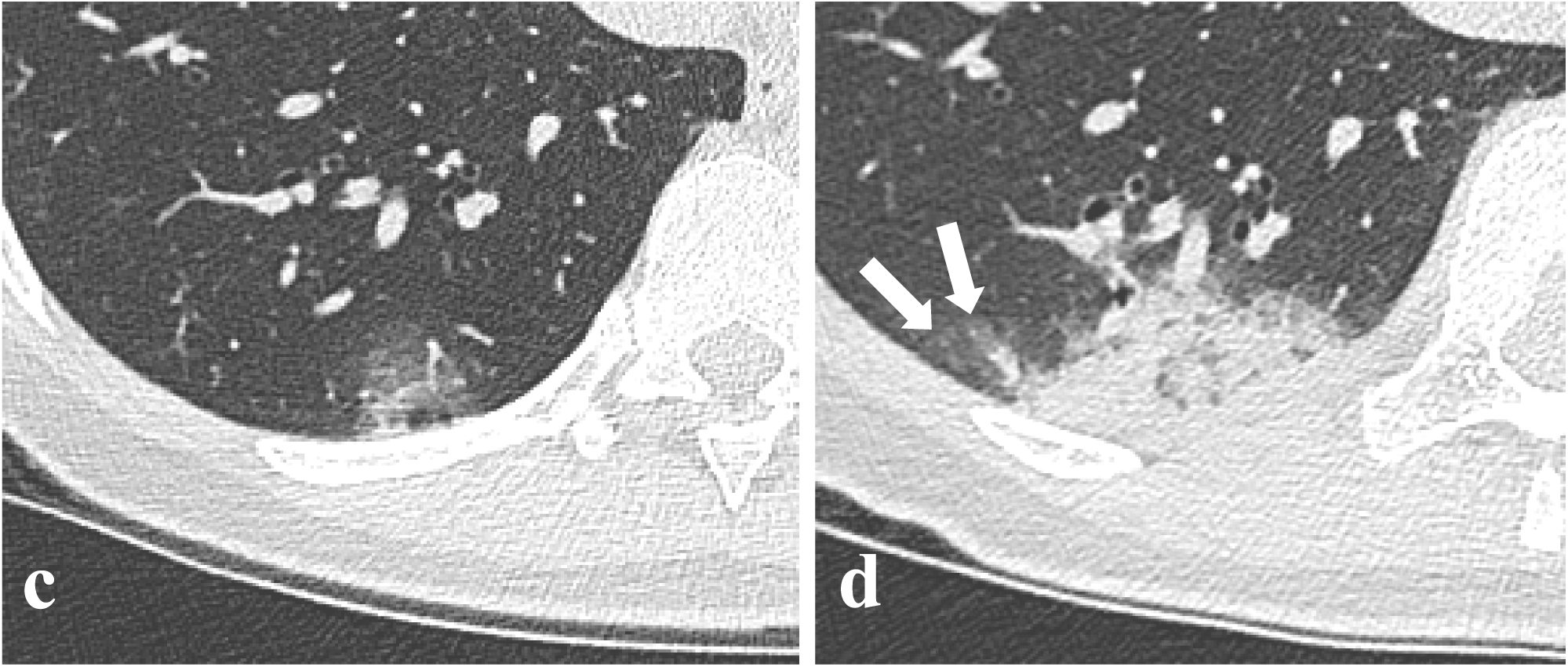
Initial CT images (a) of a 33-year-old female with COVID-19 show subpleural patchy areas of ground-glass opacity (GGO) with inter/intralobular septal thickening and air bronchogram. Follow-up CT images (b) after 5 days depict a prominent progression with an overlap pattern of GGO and consolidation. Initial (c) and follow-up (d) CT images indicate an increased volume and density of the inflammatory lesion of the right inferior lobe. Follow-up CT image also suggests the lesion with a “fried egg sign” (arrow, peripheral ground-glass halo with central solid component).

Air bronchogram sign (as a typical exhibition of pneumonia), and interlobular or intralobular septal thickening were widely observed in all cases of our study. Other common findings included roughly distinct margin (8/11, 73%), pleural retraction and thickening (7/11, 64%) with subpleural radiolucent area, intralesional vasodilatation (6/11, 55%), and a small amount of pleural effusion (1/11, 9%).

### 3.3 CT Follow-up

During the early-phase CT follow-up, the abnormal pulmonary findings show progressions in different forms in all patients. Gradually increased GGOs and consolidations with reticulations and stripes were main findings on the follow-up CT scans. In the quantitative analysis, significantly increased nV of GGO (*p* = 0.003) and consolidation (*p* = 0.003) were discovered between initial and follow-up UHR-CT (Table 3). However, the early-phase progression of each GGO lesion in our study was diverse and overlapping (including increased, enlarged, consolidated, merged, blurred and partially absorbed lesions).

## 4. Discussion

Viruses in the same family may share a similar genetic structure, pathogenesis pattern and radiological characteristics. The 2019 novel coronavirus, also known as SARS-CoV-2, is a new sub-type of subfamily *Coronavirinae* with genetic similarity to bat-like SARS coronavirus [11]. Currently, no effective treatment or reliable vaccine is available for the emerging infectious disease.

Previous reports of COVID-19 advocated the routine application of chest CT (with a sensitivity of 98%) for patients with epidemiologic suspicions but negative rRT-PCR results [9]. UHR-CT with maximum matrix size improved the spatial resolution and image quality of CT scans, which was more suitable for the assessment of GGO, intralobular reticulation, and intralesional vessels than conventional chest CT [12]. In this study, UHR-CT provided visualized details about the radiological pattern of imported early-stage COVID-19 related pneumonia.

Predominantly bilateral multifocal subpleural distributed GGOs with relatively clear margins were common imaging features of imported early-stage COVID-19 in our cohort. GGOs with marked interlobular or intralobular septal thickening, also regarded as “crazy paving sign”, was one of the most typical chest CT findings (Figure 2). Characterization of crazy paving sign may be pathologically attributable to partial alveolar filling or collapse, and thickening of interstitial tissues with increased capillary volume [13]. Chung et al. revealed a typical manifestation of COVID-19 of subpleural ground-glass lesions with consolidation [4,5,14], which is consistent with our research.

To the best of our knowledge, adjacent pleural retraction, local pleural thickening, and intralesional vasodilatation on UHR-CT were firstly reported in our study, and might be helpful for the timely screening and differential diagnosis of COVID-19. We assumed possible histological finding of focal cellular fibromyxoid exudates, granulation tissue formation of subpleural and perivascular airspaces might lead to the above radiological signs, which require further radiologic-histopathologic analysis [13,15].

On the early-phase follow-up CT images, significantly increased volume of both GGO and consolidation could be frequently observed with the infection rapidly progressed. Our findings seemed to be consistent with previous preliminary reports that follow-up CT images displayed rapid progression of pulmonary lesions with increasing number and density of GGOs and consolidations [5,6]. Moreover, we reported for the first time the characteristic “fried egg sign” of peripheral GGO halo with central solid component on follow-up CT images (Figure 3), which may help to indicate the early progress of the pneumonia. A recent study also demonstrated predominantly GGO abnormality in the early course of COVID-19, and greater consolidated areas, developed “crazy paving sign” and increased total lung involvement in the following course [7].

The majority of pulmonary GGOs in our study exhibited overlaps of imaging features and evolution modes. “Migrans” of GGOs (enlarged and absorbed lesions at the same time) could be found in some cases based on the UHR-CT images comparison (Figure 2), corroborating previous studies [6]. Analogously, another study of COVID-19 revealed that both GGOs and consolidation developed with a variety of patterns during the CT follow-up with four-day intervals [14]. Unexpectedly, our study suggested that patients with mild symptoms (as the majority of COVID-19 cases) might manifest rapid short-term progression of lung lesions even after immediate and appropriate treatment, which deserved close observation and follow-up by clinicians. More sensitive recognition and more accurate segmentation of pulmonary lesions by applying artificial intelligence technology should be further investigated in the quantitative assessment of the inflammatory lesions evolution.

Our study has several limitations. Firstly, our study was based on a short-term follow-up and a small sample from a single center. Furthermore, most COVID-19 cases in our study were imported from the remote endemic center, and lack pediatric population and severe infection. In addition, some patients may receive early medical intervention before the base-line CT scans. Finally, some fibrous lesions, incidental nodules (not related to viral inflammation), motion artifact and intralesional vessels might become confounding factors in the artificial intelligence-based analysis.

In summary, UHR-CT imaging patterns of peripleural GGO with interlobular and intralobular septal thickening, pleural retraction and thickening, and intralesional vasodilatation indicate the preliminary diagnosis of COVID-19. Most imported COVID-19 cases manifest marked progressed volume of both GGOs and consolidations during the early-phase CT follow-up. The UHR-CT based artificial intelligence methodology should be further explored for assessing pneumonia development in COVID-19 patients.

## Data Availability

The data used in the study is not publicly available.

## References

1. Chan JF-W, Yuan S, Kok K-H et al. A familial cluster of pneumonia associated with the 2019 novel coronavirus indicating person-to-person transmission: a study of a family cluster. Lancet 2020:395:514–523

2. Li Q, Guan X, Wu P et al. Early Transmission Dynamics in Wuhan, China, of Novel Coronavirus–Infected Pneumonia. New Engl J Med 2020: https://doi.org/10.1056/NEJMoa2001316

3. Antonio GE, Wong KT, Chu WCW et al. Imaging in Severe Acute Respiratory Syndrome (SARS). Clin Radiol 2003:58:825–832

4. Chung M, Bernheim A, Mei X et al. CT Imaging Features of 2019 Novel Coronavirus (2019-nCoV). Radiology 2020: https://www.doi.org/10.1148/radiol.2020200230:200230

5. Song F, Shi N, Shan F et al. Emerging Coronavirus 2019-nCoV Pneumonia. Radiology 2020: https://www.doi.org/10.1148/radiol.2020200274

6. Pan Y, Guan H, Zhou S et al. Initial CT findings and temporal changes in patients with the novel coronavirus pneumonia (2019-nCoV): a study of 63 patients in Wuhan, China. Eur Radiol 2020: https://doi.org/10.1007/s00330-020-06731-x

7. Bernheim A, Mei X, Huang M et al. Chest CT Findings in Coronavirus Disease-19 (COVID-19): Relationship to Duration of Infection. Radiology 2020: http://www.doi.org/10.1148/radiol.2020200463:200463

8. Xie X, Zhong Z, Zhao W et al. Chest CT for Typical 2019-nCoV Pneumonia: Relationship to Negative RT-PCR Testing. Radiology 2020: https://www.doi.org/10.1148/radiol.2020200343

9. Fang Y, Zhang H, Xie J et al. Sensitivity of Chest CT for COVID-19: Comparison to RT-PCR. Radiology 2020: http://www.doi.org/10.1148/radiol.2020200432:200432

10. Wang D, Hu B, Hu C et al. Clinical Characteristics of 138 Hospitalized Patients With 2019 Novel Coronavirus–Infected Pneumonia in Wuhan, China. JAMA 2020: https://doi.org/10.1001/jama.2020.1585

11. Benvenuto D, Giovanetti M, Ciccozzi A et al. The 2019-new coronavirus epidemic: Evidence for virus evolution. J Med Virol 2020: https://www.doi.org/10.1002/jmv.25688:1-5

12. Hata A, Yanagawa M, Honda O et al. Effect of Matrix Size on the Image Quality of Ultra-high-resolution CT of the Lung. Acad Radiol 2018:25:869–876

13. Franks TJ, Chong PY, Chui P et al. Lung pathology of severe acute respiratory syndrome (SARS): a study of 8 autopsy cases from Singapore. Hum Pathol 2003:34:743–748

14. Pan F, Ye T, Sun P et al. Time Course of Lung Changes On Chest CT During Recovery From 2019 Novel Coronavirus (COVID-19) Pneumonia. Radiology 2020: https://www.doi.org/10.1148/radiol.2020200370

15. Xu Z, Shi L, Wang Y et al. Pathological findings of COVID-19 associated with acute respiratory distress syndrome. Lancet Resp Med https://doi.org/10.1016/S2213-2600(20)30076-X

